# How often do physicians record the International Classification of Diseases 10^th^ Revision “Z-Codes” for social determinants of health? A nationwide analysis using the National Ambulatory Medical Care Survey

**DOI:** 10.1101/2025.04.03.25325190

**Authors:** Robert Wooten, Andrea McKinnond, Sobia S. Hussaini, Amber K. Brooks, Charles T. Jensen, Ethan Stonerook, Chris Gillette

## Abstract

**Objective:** Racial/ethnic minoritized patients and those who lack health insurance are at the highest risk for experiencing healthcare disparities, mainly because of systemic disparities in the provision of care, termed social determinants of health (SDOH). Starting with the International Classification of Diseases 10^th^ revision (ICD10), physicians can record SDOH factors that may impact their patients’ health. The objective of this study was to describe the recording of SDOH ICD10 ‘Z-codes’ in non-federal physician clinic visits and identify the characteristics of those visits.

**Methods:** This was a cross-sectional analysis of the National Ambulatory Medical Care Survey (NAMCS) in 2016 and 2018, the only two years with available ‘Z-code’ data. We used Andersen’s Behavioral Model of Health Services Utilization to select variables for analysis.

Visits for children and adults were included in the analysis. We used appropriate unweighted and weighted descriptive statistics which accounts for the complex survey design to analyze the data.

**Results:** There were a total of 23,118 unweighted visits representing 1,744,110,765 weighted visits for 2016 and 2018. In 2016, 0.21 per 100 visits (n=32 unweighted, n=1,876,542 weighted) that included a ‘Z-code’. In 2018, there were 0.08 per 100 visits (n=10 unweighted, n=681,948 weighted visits) that included a ‘Z-code’. Almost all (92.86%, n=39) physician Z-code recording was for patients who was of White race and the majority of visits with a ‘Z-code’ (59.52%, n=25) were paid for by private insurance.

**Conclusions:** Consistent with smaller-scale research, these results show that physicians may not be using ‘Z-codes’ with their racial ethnic minoritized patients, suggesting a potential disparity in recording SDOH ‘Z-codes’. More work is needed to understand methods to improve uptake of ‘Z-codes’.

## 1. Introduction

Social determinants/drivers of health (SDOH) are non-medical sociocultural factors, such as poverty, job/food insecurity, racism, homelessness, reliable transportation, etc., that negatively impact peoples’ health.^1^ It is well-known that Black, Hispanic/Latino, American Indian, and Alaska Native Americans are at high risk of experiencing SDOH disparities that impact health outcomes.^2^ Racial and ethnic minorities in the United States (US) face longstanding health disparities which primarily stem from social hierarchies and not underlying genetic differences. One of the most well-documented examples of racial health disparities reveals that Black Americans’ life expectancy, particularly Black men’, has always been lower than White Americans. In 2018, Black Americans’ life expectancy was 3.6 years shorter than non-Hispanic White Americans’.^3^ According to the Centers for Disease Control and Prevention (CDC), the most common causes of death for Black Americans are heart disease, cancer, Alzheimer’s disease, and stroke.^4^ Black Americans’ reports of why their health outcomes are poorer compared to other racial groups is the lack of high-quality medical care where they live as well as being more likely to be uninsured.^6^ This disparity in lack of access to care is more pronounced in the southern US in states that have not expanded Medicaid through the Affordable Care Act.^7–8^

In 2016, the International Classification of Diseases, 10^th^ Revision (ICD10) introduced SDOH diagnosis codes, known colloquially as ‘Z-codes’, allowing organizations and providers to document SDOH factors which negatively impact their patients’ health.^9^ Addressing SDOH has been shown to improve health behaviors and health outcomes so it is important for providers to recognize SDOH factors that their patients are experiencing.^10–11^ There has yet to be a nationally-representative examination of the use of ‘Z-codes’ in Americans without Medicare. Prior research has shown that less than 1% of visits had a Z-code being recorded; however, most of the previous research has focused on state-wide analyses or of specific populations (e.g., traditional Medicare).^12–14^ To our knowledge, there has been no investigation of SDOH factors using the National Ambulatory Medical Care Survey (NAMCS). The NAMCS was first conducted in 1973 and has been conducted each year since 1989.^15^ The purpose of this study was to describe the use of ‘Z-codes’ in non-federal physician clinic visits and identify the characteristics of those visits.

## 2. Materials and Methods

### 2.1 Study Overview

This study was approved by the Institutional Review Board (IRB) at the first authors’ institution. We conducted a cross-sectional analysis of the National Ambulatory Medical Care Survey (NAMCS) in 2016 and 2018, the only two years with available ‘Z-code’ data. The NAMCS is nationally representative of non-federal physician visits. All visits were included in the analysis because the ‘Z-codes’ can be recorded for all patients. The NAMCS survey uses a multistage probability sampling design that produces an unbiased national estimation. More information on the NAMCS survey and the estimation procedure can be found on the National Center for Health Statistics website.^16^

### 2.2 Theory and Measures

We used Andersen’s Behavioral Model of Health Services Utilization to guide variable selection.^17^ This theory states that health care utilization is a complex interplay between predisposing factors (e.g., patient age, gender, etc.), enabling factors (e.g., insurance, geographic location, availability of clinics, etc.), and need factors (e.g., perceived health status, perceived severity of the health problem). **Figure 1** presents our operationalization of Andersen’s Behavioral Model of Health Services Utilization using the available NAMCS variables.

**Figure 1:**
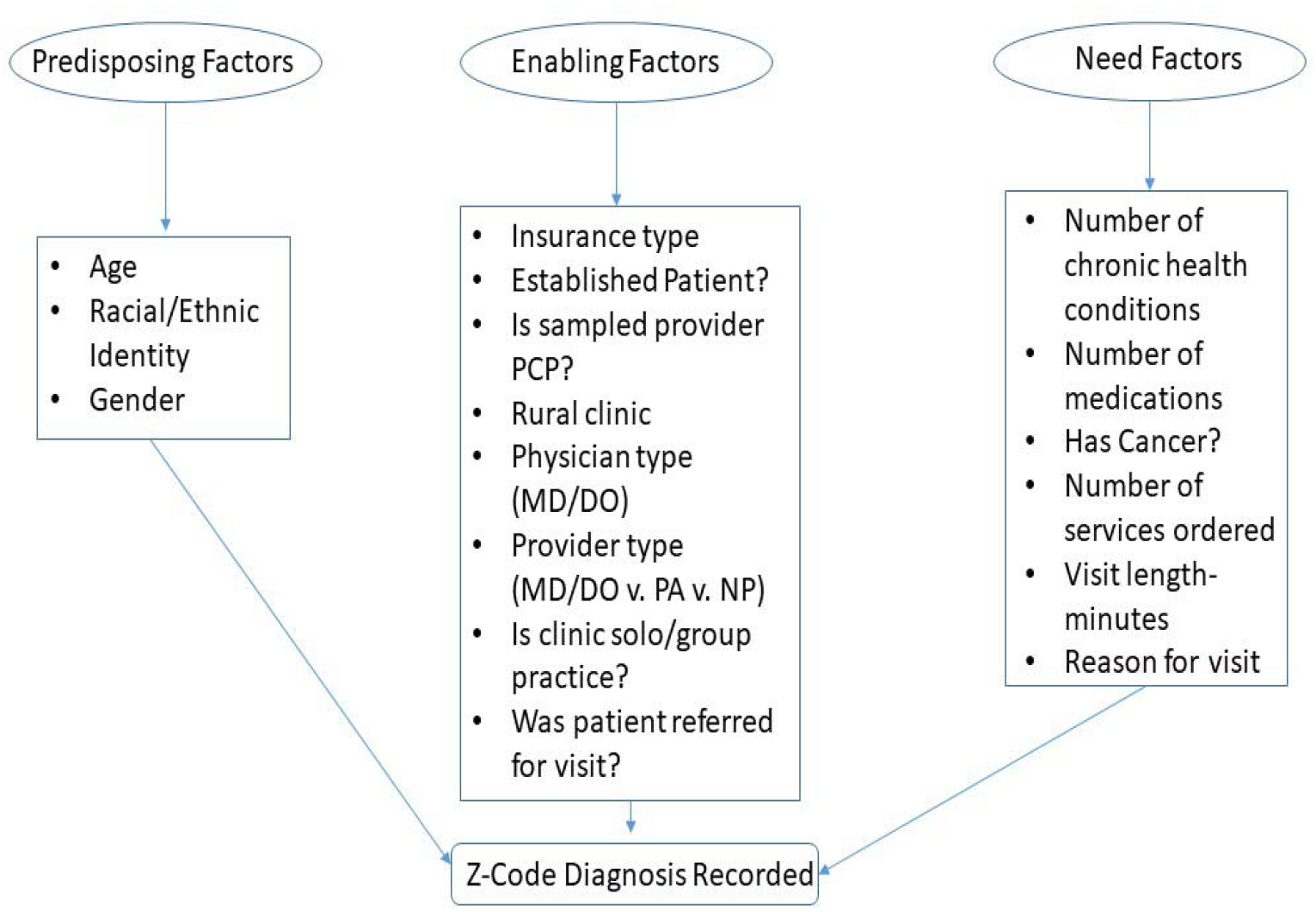
Operationalization of Andersen’s Behavioral Model of Health Services Utilization.

### 2.3 Statistical Analyses

We identified all visits in which an ICD-10 code between Z550 and Z659 were recorded by the physician in any of the five diagnosis code variables. We used appropriate unweighted and weighted descriptive statistics using PROC SURVEYFREQ and PROC SURVEYMEANS to describe the patient visits in which a physician recorded a ‘Z-code’ to account for the complex survey design. SAS v9.4 (Cary, NC) was used to analyze the data.

## 3. Results and Discussion

### 3.1 Results

**Table 1** presents the visit characteristics. In 2016, there were 13,165 unweighted visits (883,725,126 weighted visits), of which 32 unweighted visits (1,876,542 weighted visits, 0.21/100 visits) included a ‘Z-code’. In 2018, there were 9,953 unweighted visits (860,385,639 weighted visits), of which 10 unweighted visits (681,948 weighted visits, 0.08 per 100 visits) visits included a ‘Z-code’. In 2016, 31 of the 32 ‘Z-codes’ (96.88%) were recorded for patients whose race was identified as White only, 0 for Black patients, and 1 for Other (Asian). In 2018, only 1 of the 10 (90%) ‘Z-codes’ was recorded for a Black patient. **Table 2** presents all of the ‘Z-codes’ that were recorded during 2016 and 2018. The two most common ‘Z-codes’ were for ‘Other problems related to social environment’ (n=5) and ‘Problems in relationship with spouse or partner’ (n=5).

**Table 1:**
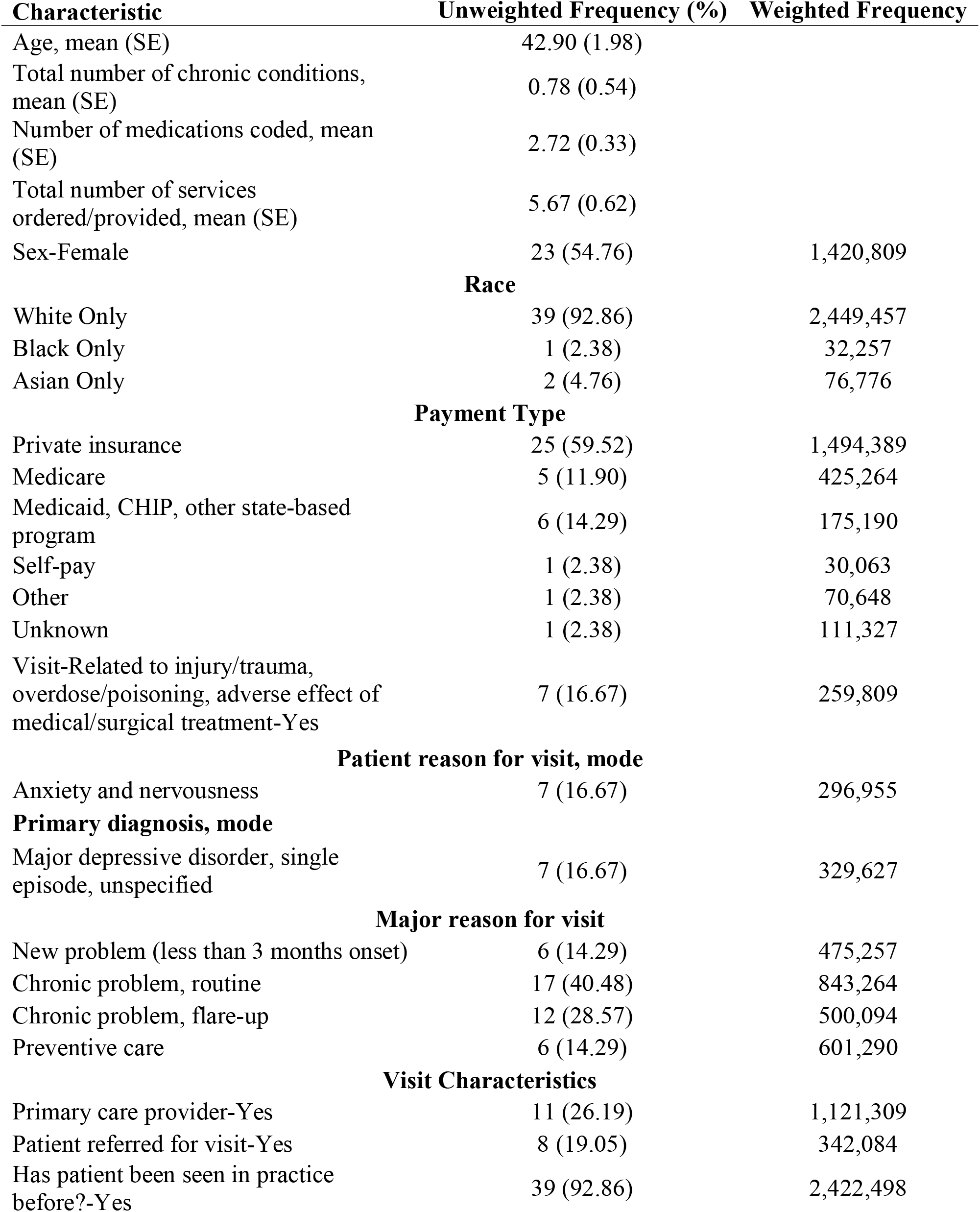
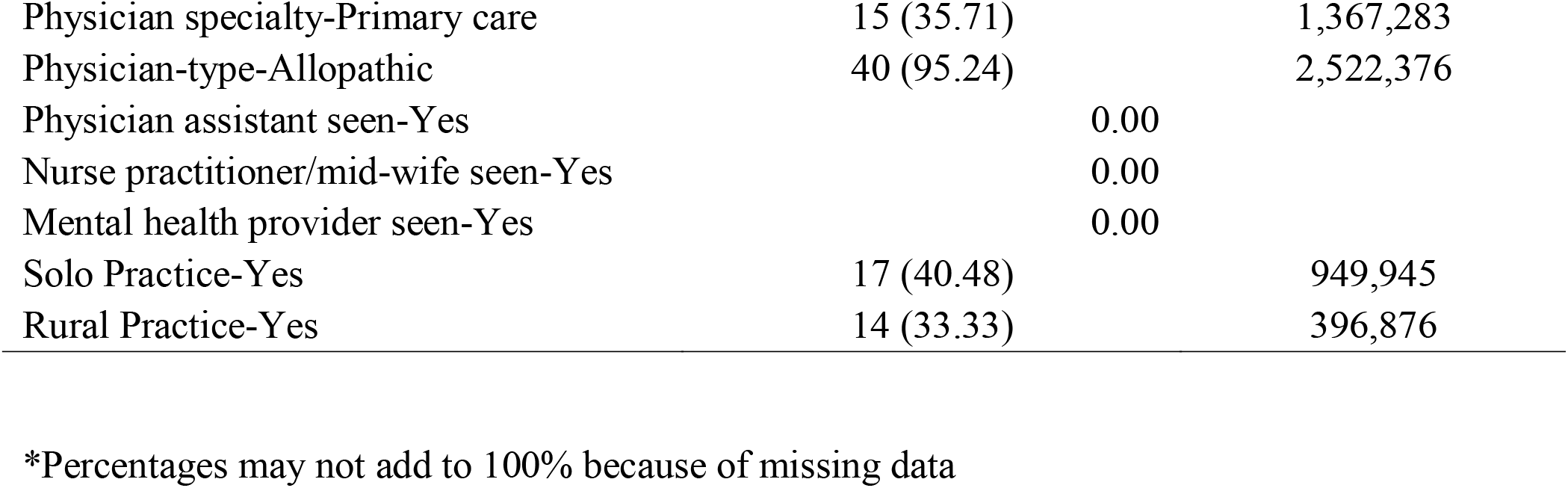
Visit Characteristics with a Z-Code (unweighted n=42, weighted n=2,558,490)

| **Characteristic** | **Unweighted Frequency (%)** | **Weighted Frequency** |
| --- | --- | --- |
| Age, mean (SE) | 42.90 (1.98) |  |
| Total number of chronic conditions, mean (SE) | 0.78 (0.54) |  |
| Number of medications coded, mean (SE) | 2.72 (0.33) |  |
| Total number of services ordered/provided, mean (SE) | 5.67 (0.62) |  |
| Sex-Female | 23 (54.76) | 1,420,809 |
| **Race** |  |  |
| White Only | 39 (92.86) | 2,449,457 |
| Black Only | 1 (2.38) | 32,257 |
| Asian Only | 2 (4.76) | 76,776 |
| **Payment Type** |  |  |
| Private insurance | 25 (59.52) | 1,494,389 |
| Medicare | 5 (11.90) | 425,264 |
| Medicaid, CHIP, other state-based program | 6 (14.29) | 175,190 |
| Self-pay | 1 (2.38) | 30,063 |
| Other | 1 (2.38) | 70,648 |
| Unknown | 1 (2.38) | 111,327 |
| Visit-Related to injury/trauma, overdose/poisoning, adverse effect of medical/surgical treatment-Yes | 7 (16.67) | 259,809 |
| **Patient reason for visit, mode** |  |  |
| Anxiety and nervousness | 7 (16.67) | 296,955 |
| **Primary diagnosis, mode** |  |  |
| Major depressive disorder, single episode, unspecified | 7 (16.67) | 329,627 |
| **Major reason for visit** |  |  |
| New problem (less than 3 months onset) | 6 (14.29) | 475,257 |
| Chronic problem, routine | 17 (40.48) | 843,264 |
| Chronic problem, flare-up | 12 (28.57) | 500,094 |
| Preventive care | 6 (14.29) | 601,290 |
| **Visit Characteristics** |  |  |
| Primary care provider-Yes | 11 (26.19) | 1,121,309 |
| Patient referred for visit-Yes | 8 (19.05) | 342,084 |
| Has patient been seen in practice before?-Yes | 39 (92.86) | 2,422,498 |

|  |  |  |  |
| --- | --- | --- | --- |
| Physician specialty-Primary care | 15 (35.71) |  | 1,367,283 |
| Physician-type-Allopathic | 40 (95.24) |  | 2,522,376 |
| Physician assistant seen-Yes |  | 0.00 |  |
| Nurse practitioner/mid-wife seen-Yes |  | 0.00 |  |
| Mental health provider seen-Yes |  | 0.00 |  |
| Solo Practice-Yes | 17 (40.48) |  | 949,945 |
| Rural Practice-Yes | 14 (33.33) |  | 396,876 |

\*Percentages may not add to 100% because of missing data

**Table 2:** All Social Determinants of Health ICD-10 Z-Codes Used.

| **Diagnosis Code Description** | **Total** |
| --- | --- |
| Other problems related to social environment | 5 |
| Problems in relationship with spouse or partner | 5 |
| Problems related to education and literacy, unspecified | 4 |
| Other problems related to upbringing | 4 |
| Other physical and mental strain related to work | 3 |
| Other problems related to psychosocial circumstances | 3 |
| Underachievement in school | 3 |
| Disappearance and death of family member | 3 |
| Problem related to social environment, unspecified | 2 |
| Disruption of family by separation and divorce | 2 |
| Problems related to other legal circumstances | 2 |
| Problem related to unspecified psychosocial circumstances | 2 |
| Occupational exposure to other risk factors | 1 |
| Problem related to housing and economic circumstances, unspecified | 1 |
| Problems related to multiparity | 1 |
| Problem related to primary support group, unspecified | 1 |

### 3.2 Discussion

It is well-known that non-White Americans, especially those who identify as Black, are at higher risk for SDOH factors negatively impacting their health outcomes.^2^ We found, using a nationally-representative dataset of non-federal physician clinic visits in the United States, that more than 9 out of every 10 visits that included a SDOH ICD10 codes were used for White patients with private health insurance. Previous research has shown that identifying and addressing SDOH factors can improve health behaviors and outcomes.^10–11^ The fact that physicians are not currently documenting SDOH for non-White Americans is indicative of a potential racial and ethnic health disparity. The ‘Z-codes’ use were low for everyone across both years, however, this is extremely problematic for non-White Americans since they are some of the most affected by SDOH factors. Additionally, those who have no insurance or who are on Medicare and/or Medicaid are also more likely to experience SDOH factors. The rates of ‘Z-code’ usage in our study are similar but lower than other studies.^12–14^ The Centers for Medicare and Medicaid study that examined ‘Z-codes’ recorded for non-elderly patients were different from our study, with the exception for spousal relationship problems, than patients who were on fee-for-service Medicare.^14^

In this study and in the Medicare study, women accounted for the majority of visits in which ‘Z-codes’ were recorded.^14^ In fact, the percentage of women who had a ‘Z-code’ recorded in the Medicare study (54.6%) was remarkably similar to our present study (54.76%).^14^ This may be attributed to ‘benevolent sexism’ (the belief that a particular gender is frail and in need of protection from harm) which typically impacts women.^18^. Another possibility is that physician judgement and heuristics in the American culture leads to an increase in ‘Z-code’ usage for women patients.^18^ Future research should investigate whether these factors or others impact physician’s’ recording of ‘Z-codes’ in certain populations.

It is also notable that there were no visits in which PAs and NPs were included that a ‘Z-code’ was recorded. We also found that allopathic physicians were responsible for almost all of the ‘Z-code’ recording nationally (95.24%). This indicates not only a potential racial/ethnic health disparity, but also a professional behavior disparity. Future research needs to identify potential reasons for these discrepancies so that more physicians, PAs, and NPs can document SDOH factors for their patients.

While the results of our study are interesting, there are limitations that affect generalizability and internal validity. First, NAMCS only collects 5 variables where the physician can document a diagnosis code, so it is possible that the physician may have recorded a ‘Z-code’ if the patient had more than 5 diagnoses. The study published by Medicare looked at all diagnoses, which may account for the differences in our findings compared to that earlier publication.^14^ Second, diagnosis coding may be impacted by the physicians’ clinic, which is not possible to identify in the NAMCS datasets. Another potential source of bias is the physicians themselves: physicians may wish to avoid perceived bias by not the ‘Z-codes’ for patients who are socio-demographically at-risk of SDOH factors. We also did not compare visits that included a ‘Z-code’ with visits that did not include a ‘Z-code’ to identify potential correlates for ‘Z-code’ recording. Future research should compare and examine these visits to identify potentially modifiable features to increase the rate of ‘Z-code’ reporting.

### 3.3 Conclusion

Improving SDOH documentation can ensure that physicians are aware of and can help mitigate the effect of these factors on Americans’ health. This study provides potential evidence of a potentially unintended healthcare disparity in that American physicians only recorded 1 SDOH ‘Z-code’ for a demographically at-risk patient while recording almost 90% of ‘Z-codes’ for White patients who have private insurance.

### 3.4 Implications

The results suggest that physicians may not be using ‘Z-codes’ with their racial/ethnic minoritized patients. This may propagate health disparities by not identifying preventable SDOH factors that are negatively impacting their patients’ health. Future research needs to investigate whether this is a true disparity or a limitation of the NAMCS datasets.

## Data Availability

All data produced in the present study are available upon reasonable request to the authors

